# Trends and risk factors associated with severity and mortality related to hepatitis A in French hospitals: a national population-based study, 2013 to 2024

**DOI:** 10.64898/2026.04.30.26351819

**Authors:** Anne-Marie Roque-Afonso, Charlotte Mouliade, Lucia Parlati, Nathalie Goutté, Julie Figoni, Samir Bouam, Philippe Sogni, Vincent Mallet

## Abstract

**Background:** As hepatitis A (HA) incidence declines in Europe and infections occur later in life, clinical presentations may worsen, particularly during outbreaks involving adults.

**Aim:** We analysed temporal trends and factors associated with severe disease and mortality among patients hospitalised for HA in France between 2013 and 2024.

**Methods:** ICD-10 codes B150 or B159 as primary discharge diagnosis were used to identify HA cases from the National Discharge Data Set. Severity (hepatic and/or extrahepatic organ failure within 12 weeks post-admission) and mortality were analysed using adjusted odds ratios in original and propensity-matched samples. Trends were assessed across five periods covering the 2017 epidemic and COVID-19, with 2013–2016 as reference.

**Results:** Among 7,928 cases (60.6% male; median age 30) 29.1% developed severe HA, and 1.43% died. Risk of severe HA increased with age (+17% of risk per decade, p < 0.001), male sex (+39%, p < 0.001), smoking (+25%, p=0.024), liver risk factors (+32%, p=0.026), and cirrhosis (+48%, p = 0.024). Risk of death increased with cirrhosis (3.55-fold, p < 0.001) and high Charlson Comorbidity Index (CCI) (9.95-fold, p < 0.001), but not with advanced age. Compared with 2013–2016, severe HA increased by 60% (p<0.001) and case fatality increased 2.22-fold (p=0.003) in 2021–2024.

**Conclusions:** HA severity and mortality have increased in France over the last decade, with advanced age and male sex increasing severity but not mortality, and high CCI limiting access to organ support, thereby increasing mortality in frail patients. Our findings highlight the need for targeted prevention and optimized care strategies for high-risk groups.

## Introduction

Hepatitis A virus (HAV) is transmitted via the faecal-oral route from person-to-person or through contaminated food or water, and therefore closely linked to socio-economic and hygiene conditions. Hepatitis A (HA) is mainly asymptomatic in children, but the frequency and severity of symptoms increase with age. In rare cases, patients develop fulminant hepatitis, which can lead to multiple organ failure with a high mortality rate, reaching up to 5% in people over 50 years of age, particularly when liver transplantation (LT) is not an option [1]. Over the past three decades, HA has ranked as the leading cause of acute viral hepatitis, with a global increase in the number of cases from 139.54 million in 1990 to over 160 million in 2021 [2, 3]. However, the age-standardised incidence tends to decrease, and there has been a dramatic decline in age-standardised disability-adjusted life years (DALY) rates and mortality rates, possibly reflecting advances in the management and treatment of organ failure [2, 4]. In most European countries, the incidence of HA is low and declining, and a growing proportion of the population is susceptible to HAV. This situation leads to repeated outbreaks, either foodborne (in particular related to imported food), or through human-to-human transmission among under-vaccinated high-risk groups, such as men who have sex with men (MSM), travellers to endemic areas, or vulnerable populations such as people who experience homelessness or people who use drugs [5, 6]. In addition, as the average age of infection shifts upward, severe illness becomes more common, and, indeed, hospitalization rates can be high during these outbreaks [7].

In France, HA surveillance since 2006 has shown a low incidence (<2/100,000 inhabitants) with a downward trend until the major epidemic of 2017, mainly affecting MSM [8], and a new upward trend from 2023 onwards (data available on https://www.santepubliquefrance.fr/maladies-et-traumatismes/hepatites-virales/hepatite-a/donnees/#tabs). Surveillance is based on mandatory reporting but is subject to underreporting [9, 10], and provides provides limited clinical data, with no information on severity or case-fatality. The objective of this study was to describe trends in the incidence of Hepatitis A in French hospitals between 2013 and 2024, and to identify demographic and clinical factors associated with severity and in-hospital mortality, based on national hospital discharge data.

## Methods

### Data Collection and Study population

Data were sourced from the Programme de Médicalisation des Systèmes d’Information (PMSI), the French National Hospital Discharge Data Set. During the observation period, the entire French population (68.4 million in 2022) had equal access to universal, tax-financed healthcare services. The database contains anonymized, standardised discharge summaries providing detailed patient demographics (age, sex as binary data, postal code of residency), primary and associated discharge diagnoses coded using the International Classification of Diseases, 10th Revision (ICD-10) [11], medical procedures, discharge dates, length of stay, and entry and discharge modes (e.g. emergency admission, transfer, discharge home, or in-hospital death). Each patient’s historical and underlying conditions were identified longitudinally by linking all hospital stays belonging to the same individual over the study period using unique anonymous identifiers [12]. The PMSI has demonstrated high validity for hard outcomes requiring hospital care. For sensitivity analyses and validation of our approach, see Supplementary Methods and [13].

As hospital discharge data are recorded for all hospitalisations in France, missingness was minimal for primary outcomes and key covariates, all identified using ICD-10 codes listed in Supplementary Table S1. Covariates included age, sex, alcohol use disorders (AUD), smoking habits, obesity, type-2 diabetes mellitus, cirrhosis, Human Immunodeficiency Virus (HIV) infection, liver risk factors, severe comorbidities assessed using the Charlson Comorbidity Index (CCI), and socio-economic deprivation measured by the French Deprivation Index (Fdep). AUD were identified in the same way as in global burden studies and previous research [13, 14]. Liver risk factors included all well identified causes of chronic liver disease (chronic hepatitis B and C, non-viral causes of chronic liver disease, i.e. congenital malformations, genetic disorders other than cystic fibrosis, autoimmune liver diseases, and disturbances in iron or copper metabolism). The CCI, widely used to predict a patient’s 1-year survival probability by quantifying the burden of comorbid conditions, assigns weighted scores to specific chronic diseases, and the sum of these weights, calculated by using developed ICD-10 coding algorithms, gives the patient’s score [15]. A score ≥3 indicates moderate to high comorbidity burden and therefore greater frailty. The Fdep was used to assess spatial socioeconomic and health inequalities, with scores divided into quintiles [16]: a score ≥Q4 indicates greater socioeconomic deprivation.

We performed complete case analyses for multivariable models, and no data imputation was performed. We identified all patients recorded with HA, i.e. with ICD-10 codes B150 or B159 as a primary or associated discharge diagnosis, between January 2013 and December 2024 (N=13,356). To increase specificity, we included only patients with HA as primary discharge code (i.e. n=7,928; 58.6%; see Figure S1, flowchart).

Data from national surveillance through mandatory reporting (available on https://www.santepubliquefrance.fr/maladies-et-traumatismes/hepatites-virales/hepatite-a/donnees/#tabs) were used to confirm the trends identified by our analysis of hospital data, namely variations in incidence, and variations in case severity, as reflected by the hospitalisation rate among reported cases, obtained by dividing the number of reported cases mentioning hospitalisation by the total number of reported cases, as a percentage.

### Outcome Measures

Our primary objective was to measure the incidence of severe HA and HA-related death over time. Severe HA was defined as hepatic and/or extrahepatic organ failure within 12 weeks of admission with the first record of HA as primary discharge diagnosis. The codes used to define severe HA are highlighted in yellow in Supplementary Table S1. These codes correspond to diagnosis codes of organ failure and to organ support therapeutic procedures, including LT. HA-related mortality was defined as death within 12 weeks of admission for HA.

### Data analysis

The primary explanatory variable was the period of observation. Periods were defined to take account of two major events that occurred during the 12 years of the study: the HAV epidemic in 2017, mainly affecting MSM [8] and the COVID-19 pandemic. The periods were: A) before the MSM outbreak (Jan. 2013-Dec. 2016), considered as reference period; B) MSM outbreak (Jan. 2017-Jun. 2018); C) after the MSM outbreak until the COVID-19 pandemic (Jul. 2018-Dec. 2019); D) COVID-19 pandemic, as defined for France, (Jan. 2020-Jun. 2021) [17]; and E) after the COVID-19 pandemic (Jul. 2021-Dec. 2024). Other explanatory variables were the key covariates referred to above.

Incidence rates per 100,000 population were calculated using population size data from the *Institut National de la Statistique et des Études Économiques* (https://www.insee.fr/fr/statistiques/1893198). Standardized rates were computed by direct standardization using the European Standard Population 2013 (ESP2013) [18] as the reference (detailed methodology provided in Supplementary Methods). Crude incidences stratified by age group, averaged between the sexes, are also presented for descriptive purposes and should be interpreted as unadjusted estimates. The same applies to the crude incidence of HA in the general population, calculated from numerators derived from mandatory reporting.

Adjusted odds ratios (aOR) from multivariable binary logistic regression models were used to measure association strengths. Variables with nominal 2-tailed p values < 0.05 were included in the multivariable models. Propensity scores were estimated considering the likelihood of severe HA or HA-related death, using all studied covariates to adjust for potential confounding factors across time periods. A full matching approach was then applied, whereby all individuals were retained and grouped into matched sets containing at least one patient from each period, with variable matching ratios, to minimize differences in covariate distributions between periods while preserving sample size [19]. Statistical tests were two-tailed with *p* < 0.05 signifying significance. Analyses were performed using R statistical software (R version 4.4.0 *Puppy Cup*).

## Results

### Patients’ characteristics and trends in the incidence of HA in hospitals, France, 2013-2024

A total of 7,928 hospitalisations with HA as primary discharge code were recorded between 2013 and 2024. Patients had a median age of 30.0 years (IQR: 18-48 years), and 60.63% were male. Risk factors for chronic liver disease were present in 12.02%, namely AUD (5.69%), cirrhosis (2.22%), and liver risk factors as defined in methods (4.11%); 12.76% had obesity or type-2 Diabetes; prevalence of HIV infection was 3.88%; 8.41% had a CCI≥3 (Table 1).

**Table 1.** Characteristics of patients hospitalized for HA across different periods, France, 2013-2024 (n=7,928 patients)

| Characteristic | Overall |  | Period A<br>[2013-2017[ |  | Period B<br>[2017-Jun. 2018[ |  | Period C<br>[Jul. 2018-2019] |  | Period D<br>[2020-Jun. 2021] |  | Period E<br>[Jul. 2021-2024] |  | p-value<sup>1</sup> |
| --- | --- | --- | --- | --- | --- | --- | --- | --- | --- | --- | --- | --- | --- |
|  | n | % | n | % | n | % | n | % | n | % | n | % |  |
| N included in analysis | 7,928 | 100 | 2,109 | 26.6 | 2,418 | 30.5 | 1,350 | 17.0 | 414 | 5.22 | 1,637 | 20.6 |  |
| Age, Median (IQR) | 30.0 (18.0, 48.0) |  | 27.0 (14.0, 48.0) |  | 34.0 (25.0, 48.0) |  | 27.0 (15.0, 46.8) |  | 38.0 (19.3, 61.0) |  | 28.0 (15.0, 48.0) |  | <0.001 |
| Male Sex | 4,807 | 60.63 | 1,095 | 51.92 | 1,843 | 76.22 | 740 | 54.81 | 193 | 46.62 | 936 | 57.18 | <0.001 |
| Smoking Habits | 635 | 8.01 | 141 | 6.69 | 257 | 10.63 | 101 | 7.48 | 37 | 8.94 | 99 | 6.05 | <0.001 |
| Alcohol Use Disorders | 451 | 5.69 | 113 | 5.36 | 186 | 7.69 | 65 | 4.81 | 22 | 5.31 | 65 | 3.97 | <0.001 |
| Obesity | 626 | 7.90 | 180 | 8.53 | 177 | 7.32 | 106 | 7.85 | 47 | 11.35 | 116 | 7.09 | 0.032 |
| Type-2 Diabetes | 385 | 4.86 | 127 | 6.02 | 92 | 3.80 | 54 | 4.00 | 29 | 7.00 | 83 | 5.07 | 0.001 |
| Liver Risk Factors | 326 | 4.11 | 107 | 5.07 | 82 | 3.39 | 57 | 4.22 | 20 | 4.83 | 60 | 3.67 | 0.049 |
| Cirrhosis | 176 | 2.22 | 61 | 2.89 | 42 | 1.74 | 27 | 2.00 | 13 | 3.14 | 33 | 2.02 | 0.056 |
| HIV Infection | 308 | 3.88 | 31 | 1.47 | 242 | 10.01 | 21 | 1.56 | 1 | 0.24 | 13 | 0.79 | <0.001 |
| CCI ≥ 3 | 667 | 8.41 | 170 | 8.06 | 93 | 3.85 | 117 | 8.67 | 88 | 21.26 | 199 | 12.16 | <0.001 |
| Fdep ≥ Q4 | 3,185 | 41.27 | 885 | 43.64 | 837 | 35.26 | 577 | 43.75 | 169 | 42.68 | 717 | 44.81 | <0.001 |
| Severe HA | 2,307 | 29.10 | 490 | 23.23 | 820 | 33.91 | 402 | 29.78 | 111 | 26.81 | 484 | 29.57 | <0.001 |
| HA-related Death | 113 | 1.43 | 25 | 1.19 | 17 | 0.70 | 15 | 1.11 | 14 | 3.38 | 42 | 2.57 | <0.001 |

**HA:** Hepatitis A. **HIV:** Human Immunodeficiency Virus; **CCI:** Charlson Comorbidity Index. **Fdep:** French deprivation index. Periods were A) before the outbreak among MSM; B) MSM outbreak; C) after outbreak until COVID-19 pandemic; D) COVID-19 pandemic; and E) after COVID-19 pandemic. Severe HA was defined as hepatic and/or extrahepatic organ failure within 12 weeks of admission.
<sup>1</sup> Kruskal-Wallis rank sum test; Pearson's Chi-squared test

The annual number of hospitalised HA cases ranged from 246 in 2020 to 1,997 in 2017 (supplementary Fig.2), and standardised incidence ranged from 0.73 to 6.49/100,000 (Fig. 1)

**Figure 1:**
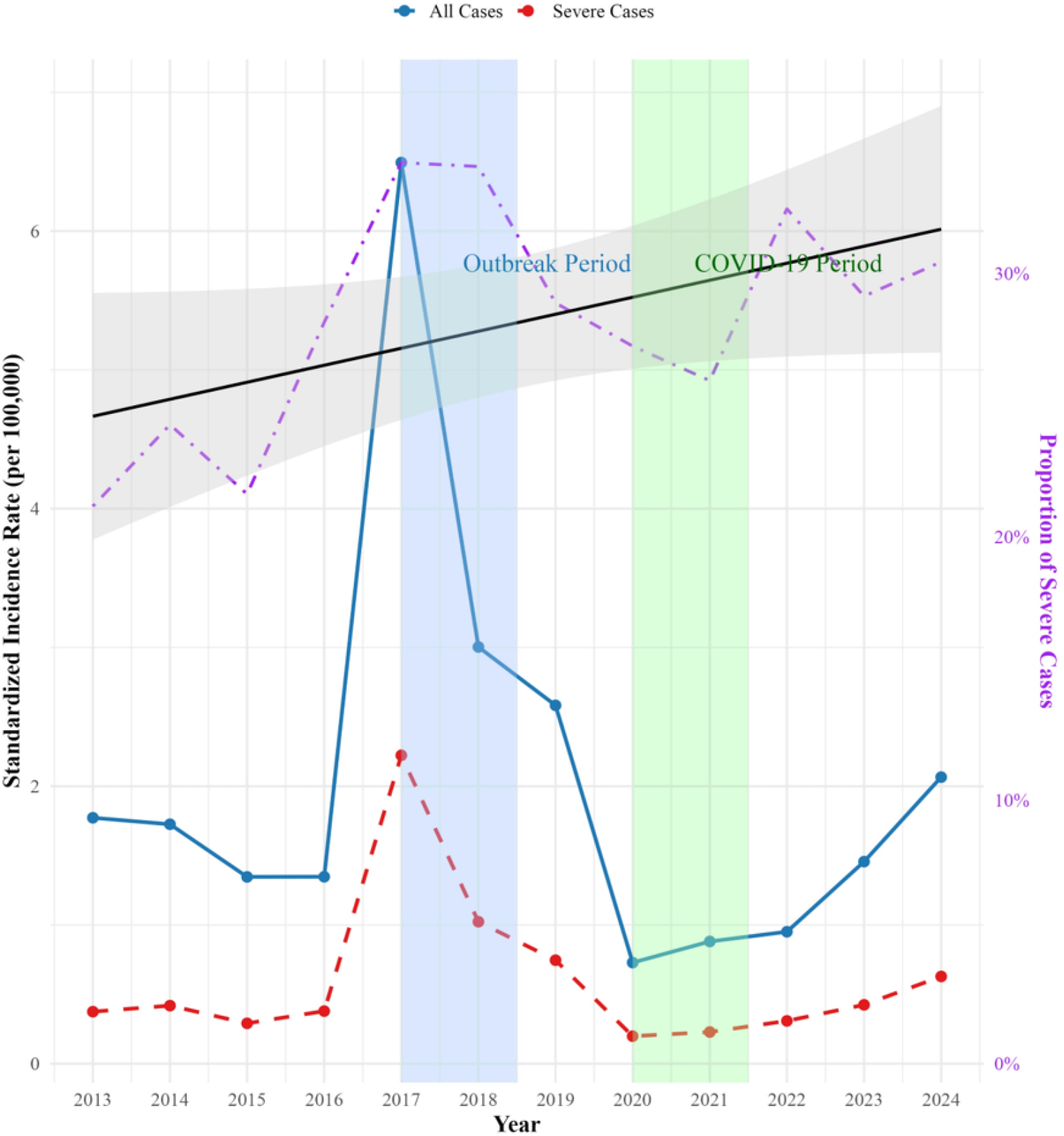
Standardised incidence of all HA cases and of severe HA cases in hospitals, per 100,000 population, France, 2013-2024. Standardized incidence rates for all HA cases (solid blue line) and for severe cases (dashed red line) are presented. The proportion of severe cases is on the right-hand axis (dotted purple line), with the black line and grey shading indicating the linear trend (p = 0.074) and its 95% confidence interval, respectively. Shaded areas indicate the MSM outbreak (blue) and the COVID-19 pandemic (green).

Incidence rates varied greatly not only from year to year but also according to sex or age group (Fig.2).

**Figure 2:**
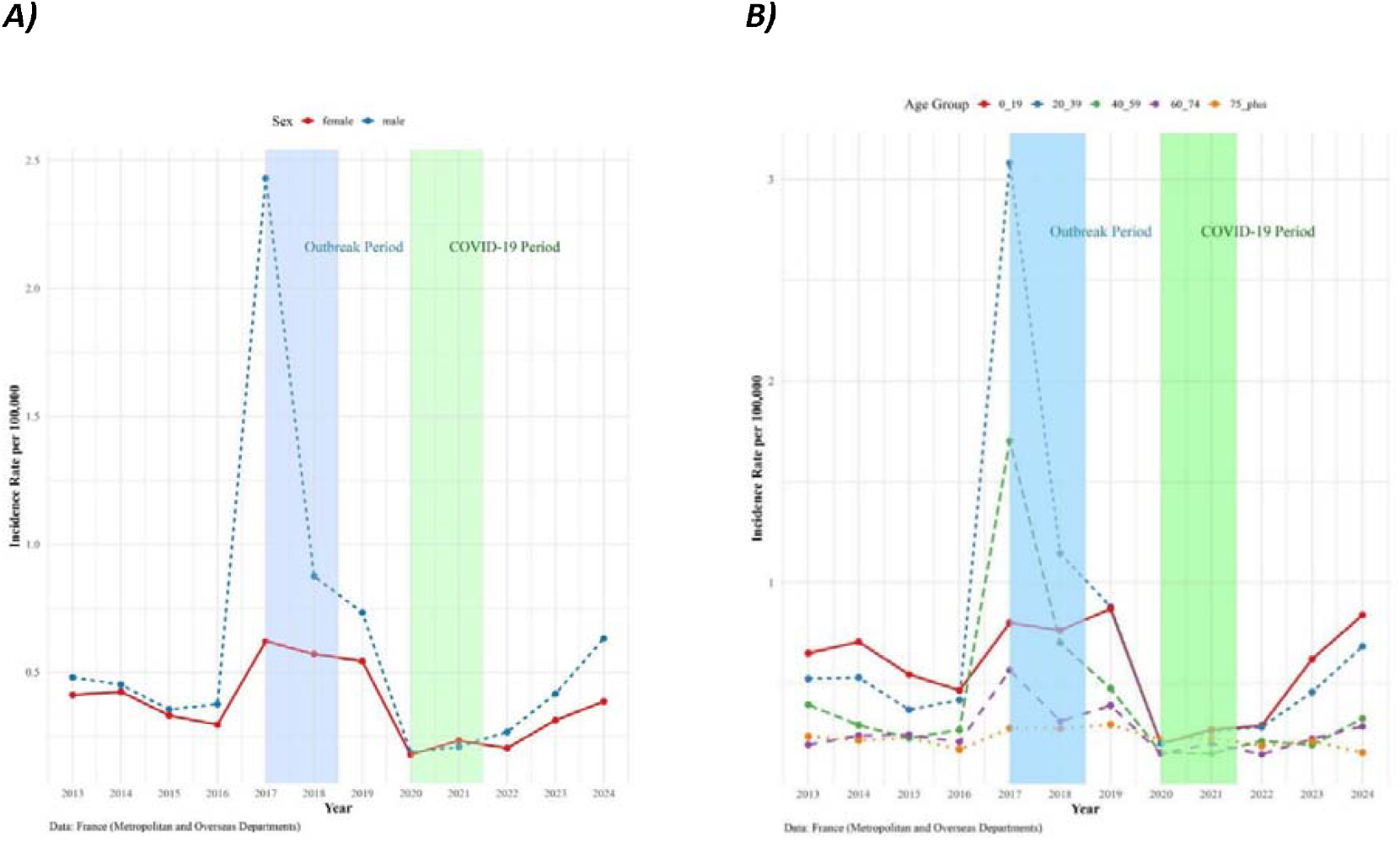
Incidence of HA by sex (A) and by age-group (B) in hospitals, per 100, 000 population, France, 2013-2024.

Patient characteristics differed particularly between B and D periods (Table 1). During period B, most patients were men (76.22%), with a high prevalence of HIV infection (10.01%), AUD (7.69%) and smoking habits (10.63%), and a low prevalence of liver risk factors (3.39%), cirrhosis (1.74%), diabetes (3.8%), obesity (7.32%), severe comorbidities (3.85%), and deprivation (35.26%). By contrast, during period D, most patients were women (53.38%), with a high prevalence of liver risk factors (4.83%), cirrhosis (3.14%), diabetes (7%), obesity (11.35%), and of CCI ≥ 3 (21.26%).

### Factors associated with severe HA in hospitals, France, 2013-2024

Severe HA was identified in 2,307 patients (29.1 %), and was associated age, sex, smoking, AUD, diabetes, liver risk factors, cirrhosis, HIV, Fdep, and the predefined study periods covering the MSM outbreak and the COVID-19 pandemic, but not obesity or CCI (Supplementary Table S2).

In a multivariate analysis, independent risk factors for severe HA (Table 2) were advanced age (+17% of risk per decade, *p* < 0.001), male sex (+39%, *p* < 0.001), smoking (+25%, *p* = 0.024), liver risk factors (+32%, *p* = 0.026), and cirrhosis (+48%, *p* = 0.024). A CCI ≥3 was negatively associated with severe HA (aOR 0.55, *p* <0.001), suggesting differences in the provision of care for patients with significant comorbidities, as the ICD-10 codes used to define severity include both diagnosis codes corresponding to organ failure (probably occurring in these frail patients) and codes corresponding to organ support therapeutic procedures. Compared with the reference period, the risk of severe HA increased by 51%, 45%, and 45% for periods B, C, and E, respectively (*p*<0.001; Table 2). A trend analysis also suggested an increase in the proportion of severe HA over time (*p*=0.074, for the trend, Fig. 2).

**Table 2:** Multivariate analysis of risks associated with severity and death in patients hospitalised for HA, France, 2013-2024.

| Characteristic | Severe HA |  | HA-related death |  |
| --- | --- | --- | --- | --- |
|  | aOR (95% CI)<sup>1</sup> | p-value | aOR (95% CI)<sup>1</sup> | p-value |
| Age, per decade | 1.17 (1.13 - 1.22) | <0.001 | 1.15 (0.98 - 1.35) | 0.089 |
| Male Sex | 1.39 (1.25 - 1.55) | <0.001 | 0.97 (0.64 - 1.47) | 0.89 |
| Smoking Habits | 1.25 (1.03 - 1.51) | 0.024 | 0.87 (0.44 - 1.63) | 0.68 |
| Alcohol Use Disorders | 1.12 (0.89 - 1.42) | 0.33 | 1.79 (0.90 - 3.42) | 0.087 |
| Obesity | 1.00 (0.82 - 1.20) | 0.97 | 1.58 (0.89 - 2.67) | 0.10 |
| Type-2 Diabetes Mellitus | 1.11 (0.87 - 1.41) | 0.41 | 0.80 (0.44 - 1.40) | 0.45 |
| Liver Risk Factors | 1.32 (1.03 - 1.68) | 0.026 | 0.56 (0.22 - 1.23) | 0.18 |
| Cirrhosis | 1.48 (1.05 - 2.08) | 0.024 | 3.55 (1.71 - 7.14) | <0.001 |
| HIV Infection | 1.17 (0.91 - 1.50) | 0.21 | 0.72 (0.11 - 2.55) | 0.66 |
| CCI ≥ 3 | 0.55 (0.43 - 0.69) | <0.001 | 9.95 (4.86 - 20.9) | <0.001 |
| Fdep ≥ Q4 | 0.92 (0.83 - 1.02) | 0.11 | 1.25 (0.84 - 1.86) | 0.27 |
| Observation period |  |  |  |  |
| A [2013-2017[ | Reference |  | Reference |  |
| B [2017-Jun. 2018[ | 1.51 (1.31 - 1.74) | <0.001 | 0.97 (0.49 - 1.87) | 0.92 |
| C [Jul. 2018-2019] | 1.45 (1.23 - 1.69) | <0.001 | 1.04 (0.52 - 2.03) | 0.91 |
| D [2020-Jun. 2021] | 1.23 (0.96 - 1.57) | 0.10 | 1.73 (0.83 - 3.51) | 0.13 |
| E [Jul. 2021-2024] | 1.45 (1.25 - 1.69) | <0.001 | 2.01 (1.18 - 3.49) | 0.011 |

<sup>1</sup> aOR = Adjusted Odds Ratio, CI = Confidence Interval.

After adjustment for all covariates using propensity score matching, the risk of severe HA increased by 49% (aOR 1.49[1.30-1.71], *p*<0.001), 54% (aOR 1.54[1.31-1.80], *p*<0.001), 36% (aOR 1.36[1.06-1.75], *p*=0.014), and 60% (aOR 1.60[1.38-1.86], *p*<0.001), during periods B, C, D and E, respectively (Supplementary Table S3 and Fig. S3-A showing adequate covariate balancing after matching).

We then examined the mandatory notification data to determine how the hospitalisation rate among reported cases had changed, hospitalisation being the marker of a particularly symptomatic or even severe infection. Mean hospitalisation rate was 51% but we observed a significant and steady increase in the proportion of hospitalised cases among those reported from 2013 onwards (*p* = 0.004, for the trend, Fig. 3).

**Figure 3:**
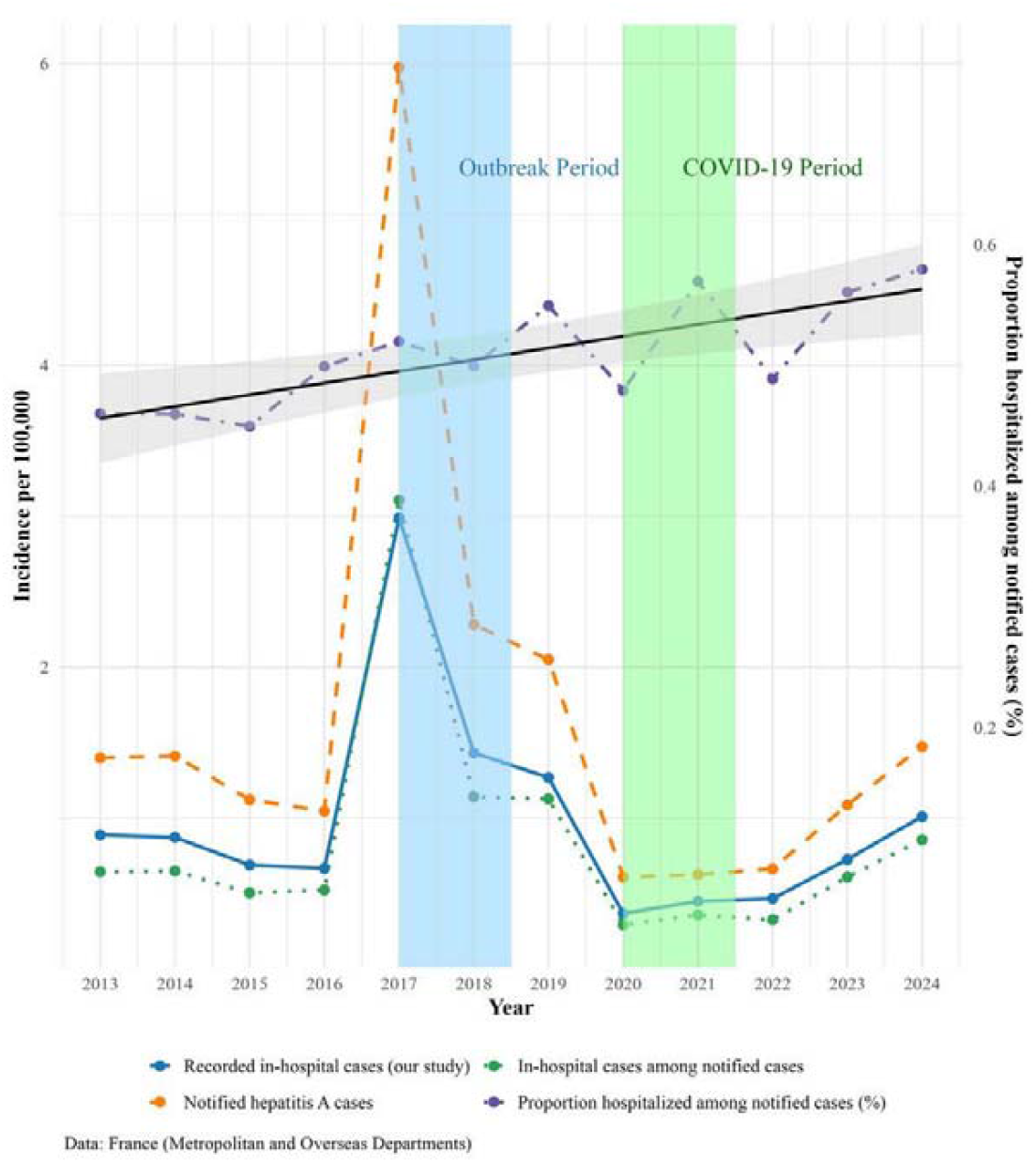
HA in the general population (mandatory reporting data), and in hospitals (National Hospital Discharge Data Set), Crude incidence per 100,000 population, France, 2013-2024. Data from mandatory reporting include incidence of notified HA cases (dashed orange line), of hospitalisation among notified cases (dotted green line), proportion of hospitalised cases among notified cases (dotted/dashed purple line), and the linear trend of hospitalisation proportion among notified cases (black line; p = 0.004). Incidence of HA cases in French hospitals (blue line) shows similar trends to other incidences.

### Factors associated with HA-related death in hospitals, France, 2013-2024

In-hospital death was recorded for 113 patients, corresponding to an overall case-fatality rate of 1.43% (Table 1). Factors associated with death age, AUD, obesity, diabetes, cirrhosis, CCI, and predefined study periods but not sex, liver risk factors, HIV, or Fdep (Supplementary Table 2).

In a multivariate analysis (Table 2), independent risk factors for death were cirrhosis (aOR 3.55, *p* < 0.001) and CCI ≥3 (aOR 9.95, *p* < 0.001). The risk of death did not vary significantly during the first four periods. However, compared to the reference period, this risk was twice as high in 2021–2024 (aOR 2.01, *p*=0.011) (Table 2). This result was confirmed after propensity score matching (supplementary Table S3 and Fig. S3-B): After adjustment for all covariates, the probability of death in 2021–2024 remained more than twice that of reference period (aOR 2.22[1.32-3.84], *p*=0.003).

## Discussion

Our retrospective study of nearly 8,000 HA-related hospitalisations in France over 12 years covered two major events: the MSM epidemic in 2017–2018 [8], when incidence of HA in hospitals peaked, and the COVID-19 pandemic in 2020–2021 [17], when it was lowest. Overall, 29.1% of hospitalised patients developed severe HA, and 1.43% died. This case fatality rate is consistent with the 1.30% rate reported in France during 2008–2013 [20]. As others have noted, determinants of severity and mortality differ [21-25]. In our cohort, each decade of age, male sex, chronic liver disease, cirrhosis, and smoking increased severity risk, while cirrhosis and CCI>3—but not age—predicted death. The latter finding likely reflects limitation on the use of organ support procedures in patients with poor physiological condition [15], as France has equitable hospital access and no link was observed between social vulnerability and outcomes. Strikingly, we observed an increased proportion of severe cases and a doubling of hospital mortality from mid-2021 onwards compared with pre-2017. This worsening of HA presentation in recent years is also reflected by the rise in hospitalisation rate among cases reported through mandatory notification. To our knowledge, such progression had not been documented previously.

Previous studies in high-income countries have analysed HA-related hospitalisations and their evolution over time. Those performed from the late 1990s to mid-2010s—periods of low and declining HA incidence in the United States, Taiwan, and Europe—reported decreasing incidence and no increase in severity or mortality, despite rising age and comorbidities [21-23, 26]. More recent studies, including ours, spanned major epidemics. An American study (1998–2020) [24] covered outbreaks among people who use drugs and people who experience homelessness from 2016 onward, and a Spanish study (2000–2021) [25] included the 2016–2018 European MSM epidemic [8]. Both reported sharp increases in incidence during epidemics, followed by declines during the COVID-19 pandemic. The U.S. data showed no significant change in hospital mortality, averaging 2.7% (23) despite high hospitalisation frequency (84.8%) and variable case fatality (0–10.8%) among outbreak cases [27]. The Spanish study reported no trend analysis but a low mortality rate (0.3%), likely reflecting the MSM epidemics, in which cases presented very low case fatality (0.03–0.26%) [7]. Neither study analysed changes in severity, or patient characteristics across epidemic and non-epidemic periods. In France, surveillance data based on mandatory reporting have shown substantial variation in age and sex distribution depending on viral circulation and epidemic context. Accordingly, for temporal trends analysis, we defined time frames accounting for the MSM outbreak and the COVID-19 pandemic, and confirm significant differences in age, sex, and comorbidities between periods but also in case fatality, lowest in 2017–2018 (0.7%), consistent with MSM epidemic data [7], and highest during COVID-19 (3.38%), possibly reflecting the pressure on the healthcare system, as also suggested by the U.S. study [24]. Regarding clinical severity, comparison between studies is challenging because definitions vary. We defined severity by ICD-10 codes corresponding to diagnoses of organ failures and therapeutic procedures of organ support, including LT—an outcome-based, objective measure, validated in previous studies, that better reflects actual severity [13, 28]. Indicators such as prolonged hospital stay, often used as proxies, may reflect specific organisational factors [23] and are thus less reliable.

The incidence was higher among men than women throughout the study, particularly in 2017–2018, when it was more than four times higher (Fig. 2A). This period also marked a shift in the age group with the highest incidence from 0–19 to 20–39 years (Fig. 2B), consistent with MSM epidemic data showing a median age of 35 years [7]. It is likely that this shift towards older ages contributed to the highest severity rate of all periods, i.e. 34%, because most comorbidities, particularly liver disease and cirrhosis, were less common than in other periods. Only smoking and HIV infection were more common. In our study, HIV infection did not increase HA severity or mortality, consistent with findings from a large cohort showing a prolonged but milder course [29]. By contrast, smoking correlated with severity but not with death. Smoking is strongly linked to poor outcomes in chronic liver disease [30] and, although hepatitis A is self-limiting, smoking-related immune impairment could heighten risk of organ failure. Probably because young, otherwise healthy smokers can recover from liver failure with appropriate management, we did not find an association with increased mortality risk. However, unlike Wasuwanich et al., smoking was not associated with reduced mortality [24].

A major finding was the apparent worsening of HA severity over time, evidenced by the increase in hospitalisation rates among notified cases from 2013 onwards and higher severity among hospitalised cases in recent years, coinciding with resurgence in viral circulation, linked to renewed travel and loosening of hygiene measures after COVID-19. Compared with pre-MSM epidemic years, in recent years, more men and patients with severe comorbidities were affected, two risk factors associated with severity and death, respectively. But after propensity score matching for confounders, risk of severe HA and death remained higher. One hypothesis is changes in viral virulence, as proposed for recent U.S. epidemics [24, 31]. HAV does have virulence factors in genomic regions responsible for cell culture adaptation and attenuation, used in live vaccine development [32]. Some studies suggest nucleotide variations or specific genotypes may correlate with higher aminotransferase levels [1], though this does not equate to clinical severity. Given HAV’s non-cytopathic nature and complex immune-mediated pathogenesis [33], viral genetics likely plays a modest role compared with host factors. The French Reference Centre surveillance revealed no major genotype shifts and high strain diversity, arguing against emergence of a virulent variant. The observed increase in severity and mortality thus remains unexplained and warrants further investigation, including potential cognitive biases in clinical decision-making [34]. It is conceivable that, as during the COVID-19 period, care resources became more concentrated on younger MSM—perceived at higher risk—while those with multiple comorbidities received less intensive management.

Administrative databases provide large, multicentric, and diverse cohorts but have limitations. Firstly, outpatient cases are not captured, so findings apply to hospitalised patients only. Secondly, reliance on coded diagnoses and procedures may lack both sensitivity and specificity, as their accuracy depends on clinicians’ coding practices. HA diagnosis, based on anti-HAV IgM detection, is generally straightforward but still subject to miscoding or false-positive IgM results, potentially inflating case counts [23, 35]. To improve specificity, we excluded patients where HA appeared as an associated diagnosis although some with jaundice, acute liver failure or decompensated cirrhosis may have been true cases. However, our trends and case numbers aligned with national surveillance data, validating our selection approach. In addition, coding of our chosen outcomes (mortality and severity) has demonstrated 100% accuracy in prior French studies [13, 28]. Therefore, we ensured high specificity, with possible loss of sensitivity due to patients’ selection.

Safe, highly immunogenic vaccines have been available since the early 1990s, providing both pre- and post-exposure protection. In France, consistent with World Health Organisation guidance for low-endemicity areas, vaccination targets groups at increased infection risk (travellers, MSM) or severe-disease risk (chronic liver disease) [36]. French policy also includes children of immigrant parents and patients with cystic fibrosis (https://sante.gouv.fr/prevention-en-sante/preserver-sa-sante/vaccination/calendrier-vaccinal). However, it excludes migrants, prisoners, people who experience homelessness, and people who use drugs, among whom seroprevalence is decreasing, as in the general population [37]. Moreover, outbreaks among MSM demonstrated under-vaccination even within targeted groups [38]. Declining seroprevalence and recurring adult epidemics among adults argue for strengthening the current targeted strategy and possibly extending vaccination beyond high-risk groups to the general population.

## Conclusion

This national study provides the first comprehensive analysis of HA prognosis among hospitalised patients in France, where universal access minimises selection bias. Age, male sex, chronic liver disease, cirrhosis, and smoking increased the risk of severe HA, while cirrhosis and high CCI, but not age, predicted HA-related death, likely due to limited management of frail patients. Our observation of increased severity and mortality related to HA in recent years warrants confirmation in other low-endemicity countries to inform future vaccination strategies, potentially supporting broader or universal vaccination policies.

## Supporting information

Supplementary methods and results

## Statements

### Data availability

The data underlying this study are not publicly available, as access to the French National Hospital Discharge Database (PMSI) is restricted by law.

### Ethical statement

The study was approved by the *Commission nationale de l’informatique et des libertés* under registration number DR-2017-404. We used de-identified data, and informed consent was not required.

