## Supplementary methods and results for "Trends and risk factors associated with severity and mortality related to hepatitis A in French hospitals: a national population-based study, 2013 to 2024"

**Contents:**

**Supplementary Methods**

**Supplementary Tables 1-3**

**Supplementary Figures 1-3**

**Supplementary Methods**

*Data Collection and Study population*: sensitivity analyses and validation of our approach

The French Hospital Discharge Database coding system has demonstrated high validity for hard outcomes requiring hospital care. In a previously published nationwide study using the same database and analytical framework, liver-related outcomes were defined as hospital-managed events, including decompensated cirrhosis, primary liver cancer related to cirrhosis, and liver transplantation, the latter being exhaustively identified through the national transplantation registry. Mortality was assessed using in-hospital death records. In that study, a sensitivity analysis was conducted to estimate mortality occurring outside hospital, based on the morbidity profile of patients discharged with chronic liver disease and subsequently lost to follow-up. This analysis showed that in-hospital deaths accounted for 97.0% of overall mortality (95% CI: 96.6–97.3). Analyses using overall mortality instead of in-hospital mortality yielded similar results, supporting the use of in-hospital mortality and hospital-recorded liver-related outcomes as reliable endpoints in this database [Schwarzinger et al. 2017].

*Data analysis:* calculation of standardized incidence rates

Standardized rates were computed by direct standardization using the European Standard Population 2013 (ESP2013) [18] as the reference, with five age groups - 0–19, 20–39, 40–59, 60–74, and 75 years or older - weighted at 21.4%, 27.2%, 29.1%, 14.9%, and 7.4%, respectively. As ESP2013 originally specifies finer age bands, weights were aggregated to match the age strata available in the PMSI data and rescaled to sum to unity. For each calendar year, stratum-specific crude rates were calculated separately for each sex-by-age-group combination, then multiplied by the corresponding ESP2013 weight and summed to yield the annual age-sex standardized incidence rate. Severe HA standardized rates were derived using the identical procedure, restricted to hospitalizations meeting the severity definition.

**Supplementary Tables**

**Table S1: ICD10 code dictionary.**

Highlighted in yellow are the codes used to define severe Hepatitis A: i.e. hepatic and/or extrahepatic organ failure-within 12 weeks of admission with the first record of HA as primary discharge diagnosis. These codes correspond to diagnosis codes of organ failure and to organ support therapeutic procedures, including LT.

| **Diagnosis** | **ICD-10/Medical Procedure Codes** |
|---|---|
| Acute and Subacute Hepatic Failure | K72.0, K71.2 |
| Acute Hepatitis A with Hepatic Coma | B15.0 |
| Acute Hepatitis A without Hepatic Coma | B15.9 |
| Acute Kidney Injury | N17-, JVJF002, JVJF003, JVJF005, JVJF006, JVJF007, JVJF005, JVJB002 |
| Acute Respiratory Failure | J96.0, J80-, J81-, GLLD002, GLLD003, GLLD017, GLLD019, GLLD004, GLLD005, GLLD006, GLLD007, GLLD008, GLLD009, GLLD012, GLLD013, GLLD015, DKMD001, GEPA004, GELD002, GELD004, ECCO2R, GLJF010 |
| AIDS | B20-, B21-, B22-, B24- |
| Alcohol Rehabilitation | Z50.2, Z71.4 |
| Alcohol Use Disorders | E24.4, E51.1, F10.1, F10.2, F10.3, F10.4, F10.5, F10.6, F10.7, F10.8, F10.9, F10.2, F10.2, F10.2, F10.2, G31.2, G62.1, G72.1, I42.6, K29.2, K70-, K85.2, K86.0, O35.4, Z50.2, Z71.4, Z72.1 |
| Alcoholic Cirrhosis | K70.3 |
| Alcoholic Hepatic Failure | K70.4 |
| Alcoholic Hepatitis | K70.1 |
| Alcoholic Liver Disease | K70- |
| Ascitis | R18-, HPHB003, HPJB001 |
| Aspergillosis | B44- |
| Autoimmune Hepatitis | K75.4 |
| Bacteremia | A49- |

| **Diagnosis** | **ICD-10/Medical Procedure Codes** |
|---|---|
| Bronchopneumonia | J18- |
| Budd Chiari Syndrome | I82.0 |
| Cerebral Infarction | I63- |
| Cerebrovascular Disease | G45-, G46-, H34.0, I6 |
| Chemotherapy for Cancer | Z51.1 |
| Chemotherapy for Connective Tissue Disorder | Z51.2 |
| Cholangitis | K83.0, K83.1, K83.5, K83.5, K87.0, R17- |
| Chronic Hepatitis B | B18.1, B16.2, B16.9 |
| Chronic Hepatitis C | B18.2 |
| Chronic Hepatitis D | B16.0, B16.1, B18.0, B17.0 |
| Chronic Kidney Disease Advanced | I12.0, I13.1, N00-, N01-, N03-, N05-, N18.3, N18.4, N18.5, N19-, N25.0, Z49.0, Z49.1, Z49.2, Z94.0, Z99.2, HGPC005, HPGA001, HPJP001, HPKA002, HPKB001, HPKC014, HPLA005, HPLB004, HPLC035, HPPA004, HPPP002, JVRP007, JVRP008, JAEA003, JVJB001, JVRP004, JVJF004, JVJF008, JVQF001, JVQF007, JVQP002, JVQP009, YYYY007 |
| Chronic Kidney Disease | E10.2, E11.2, E12.2, E13.2, E14.2, I15.1, JAHB001, JAHH002, JAHC001, JAHA001, JAHJ006, JAHJ007, N02-, N04-, N06-, N07-, N08-, N18.1, N18.2, N19-, N25-, N08.3 |
| Chronic Obstructive Pulmonary Disease | I27.8, I27.9, J40-, J41-, J42-, J43-, J44-, J45-, J46-, J47-, J60-, J61-, J62-, J63-, J64-, J65-, J66-, J67-, J68.4, J70.1, J70.3 |
| Cirrhosis | I85.9, I86.4, I98.2, I98.2, K70.3, K71.7, K74.3, K74.4, K74.5, K74.6, K76.6 |
| Congestive Heart Failure | I09.9, I11.0, I13.0, I13.2, I25.5, I42.0, I42.5, I42.6, I42.7, I42.8, I42.9, I43-, I50-, P29.0 |
| Connective Tissue Disorder | M05-, M06-, M31.5, M32-, M33-, M34-, M35.1, M35.3, M36.0 |
| Cryptococcosis | B45.0, B45.1, B45.3, B45.7 |
| Decompensated Cirrhosis | R17-, K70.4, K71.1, K72-, K76.7, I28.0, I85.0, I98.3, I98.2, R18-, J94.8, K65-, EHBD001, EHNE002, HESE001, HESE002, HPHB003, HPJB001, EHCA003, EHCA006, EHCA009, EHCA007, EHCA004, EHCA002, EHCA005, EHCA010, EHCA001, HEPA005, HEPA004, HEPA007 |

| Diagnosis | ICD-10/Medical Procedure Codes |
| --- | --- |
| Dementia | F00-, F01-, F02-, F03-, F05.1, G30-, G31.1 |
| Diabetes Mellitus Complicated | E10.2, E10.3, E10.4, E10.5, E10.7, E11.2, E11.3, E11.4, E11.5, E11.7, E12.2, E12.3, E12.4, E12.5, E12.7, E13.2, E13.3, E13.4, E13.5, E13.7, E14.2, E14.3, E14.4, E14.5, E14.7, H36.0, N08.3, H28.0, G63.2 |
| Diabetes Mellitus Uncomplicated | E10.0, E10.1, E10.6, E10.8, E10.9, E11.0, E11.1, E11.6, E11.8, E11.9, E12.0, E12.1, E12.6, E12.8, E12.9, E13.0, E13.1, E13.6, E13.8, E13.9, E14.0, E14.1, E14.6, E14.8, E14.9 |
| Diabetes Mellitus | E1 |
| Disseminated Intravascular Coagulation | D65-, D68.4 |
| Extracorporeal Co 2 Removal | ECCO2R, GLJF010 |
| Embolism and Thrombosis of Renal Vein | I82.8 |
| Embolism and Thrombosis of Unspecified Vein | I82.9 |
| Embolism and Thrombosis of Vena Cava | I82.2 |
| Gastro Esophageal Varices Bleeding | I85.0, I98.3, I98.2, EHBD001, EHNE002, HESE001, HESE002 |
| Gastro Esophageal Varices not Bleeding | I85.9, I86.4, I98.2, I98.2 |
| Hemiplegia | G04.1, G11.4, G80.1, G80.2, G81-, G82-, G83.0, G83.1, G83.2, G83.3, G83.4, G83.9 |
| Hepatic Encephalopathy | K72.0, G94.3, R40.2 |
| Hepatocellular Carcinoma Treatment | YYYY170, EDLF017, EDLF016, HLNM001, EDQH006, EDLF014, YYYY210, HLHJ005, EDQH007, HLHJ006, EDLF015, EDQH008, HLNK001, EDSF006, EDLL001, ZZQA002, EHSF001, HLQX004, EDLL002, HLFA020 |
| Hepatocellular Carcinoma | C22.0 |
| Hepatopulmonary Syndrome | I28.0 |
| Hepatorenal Syndrome | K76.7 |
| HIV Infection | Z21-, B20-, B21-, B22-, B23-, B24- |

| Diagnosis | ICD-10/Medical Procedure Codes |
|---|---|
| Hydrothorax | J94.8 |
| Hypercholesterolemia Pure | E87.0 |
| Hyperlipemia Mixed | E78.2 |
| Hyperlipemia | E78.0, E78.1, E78.2, E78.4, E78.5 |
| Hypertension | I10-, I11-, I12-, I13-, R03.0 |
| Hypertensive Heart Failure with or without Renal Failure | I10-, I13.0, I13.2 |
| Hypertriglyceridemia | E87.1 |
| Intrahepatic Bile Duct Carcinoma | C22.1 |
| Invasive Candidiasis | B37.1, B37.5, B37.6, B37.7 |
| Jaundice | R17- |
| Leukemia | C91-, C92-, C93-, C94-, C95-, C96- |
| Liver Biopsy | HLHB001, HLHJ003, HLHJ006, HLHH006, HLHJ005, HLHH007, HLHH001, HLHH005 |
| Liver Disease Mild | K71.3, K71.4, K71.5, K73-, K74.0, K74.1, K74.2, K74.3, K76.0, K76.4, K76.5 |
| Liver Disease Moderate to Severe | K70.3, I85.0, I85.9, I86.4, I98.2, J94.8, K70.3, K70.4, K71.1, K71.7, K72.1, K72.9, K74.4, K74.5, K74.6, K65-, K66-, K76.7, R17-, R18-, EHBD001, EHNE002, HESE001, HESE002, HPHB003, HPJB001, EHCA003, EHCA006, EHCA009, EHCA007, EHCA004, EHCA002, EHCA005, EHCA010, EHCA001, HEPA005, HEPA004, HEPA007 |
| Liver Transplantation | HLEA001, HGEA002, HLEA002, HGEA004 |
| Lymphoma | C81-, C82-, C83-, C84-, C85-, C86-, C87-, C89-, C90- |
| Mechanical Ventilation | GLLD004, GLLD005, GLLD006, GLLD007, GLLD008, GLLD009, GLLD012, GLLD013, GLLD015, DKMD001, GEPA004, GELD002, GELD004 |
| Myocardial Infarction | I21-, I22-, I25.2 |
| Obesity | E66- |
| Other Cause of Chronic Liver Disease | E83.0, E83.1, FEJF003, E84.8, I82.0, K83.0, K74.3, K75.4, Q44.2, Q44.3, Q44.6 |

| **Diagnosis** | **ICD-10/Medical Procedure Codes** |
|---|---|
| Other Diabetes Mellitus | E12-, E13-, E14- |
| Other Primary Liver Cancer | C22.2, C22.3, C22.4, C22.7, C22.9 |
| Oxygen Therapy | GLLD002, GLLD003, GLLD017, GLLD019 |
| Palliative Care | Z51.5 |
| Paraplegia Hemiplegia | G04.1, G11.4, G80.1, G80.2, G81-, G82-, G83.0, G83.1, G83.2, G83.3, G83.4, G83.9 |
| Peptic Ulcer Disease | K25-, K26-, K27-, K28- |
| Peritonitis | K65- |
| Peripheral Vascular Disease | I70-, I71-, I73.1, I73.8, I73.9, I77.1, I79.0, I79.2, K55.1, K55.8, K55.9, R02-, Z95.8, Z95.9 |
| Phlebitis and Thrombophlebitis | I80- |
| Pneumocystosis | B59- |
| Pneumonia Bacterial | J13-, J14-, J15-, J16-, J17.0, J17.8, J18-, J85- |
| Pneumonia Due to Streptococcus Pneumoniae | J13- |
| Pneumonia Due to Haemophilus Influenzae | J14- |
| Pneumonia Bacterial not Elsewhere Classified | J15- |
| Pneumonia Due to Other Infectious Organisms not Elsewhere Classified | J16- |
| Pneumonia in Diseases Classified Elsewhere | J17.0, J17.8 |
| Portal Hypertension | K76.6 |
| Portal Vein Thrombosis | I81- |
| Renal Replacement Therapy | JVJF002, JVJF003, JVJF005, JVJF006, JVJF007, JVJF005, JVJB002 |
| Renal Transplantation | T86.1, Z94.0, JAEA003 |
| Shock | I46-, R57-, A48.3, EQLF001, EQLF003 |

| **Diagnosis** | **ICD-10/Medical Procedure Codes** |
| --- | --- |
| Systemic Inflammatory Response Syndrome of Infectious Origin | A40-, A41-, A49-, R57.2, R65.0 |
| Smoking | F17-, Z71.6, Z72.0, T65.2 |
| Solid Tumor Localised without Liver | C0, C1, C20-, C21-, C23-, C25-, C26-, C3, C40-, C41-, C43-, C45-, C46-, C47-, C48-, C49-, C5, C6, C70-, C71-, C72-, C73-, C74-, C75-, C76- |
| Solid Tumor Localised | C0, C1, C20-, C21-, C22-, C23-, C24-, C25-, C26-, C3, C40-, C41-, C43-, C45-, C46-, C47-, C48-, C49-, C5, C6, C70-, C71-, C72-, C73-, C74-, C75-, C76- |
| Solid Tumor Metastatic | C77-, C78-, C79-, C80- |
| Splanchnic Thrombophlebitis | I81-, I82- |
| Surgical Shunt | EHCA003, EHCA006, EHCA009, EHCA007, EHCA004, EHCA002, EHCA005, EHCA010, EHCA001, HEPA005, HEPA004, HEPA007 |
| Syphilis | A51-, A52-, A53- |
| Thrombophlebitis Migrans | I82.1 |
| Transfusion | FELF003, FELF004 |
| Transjugular Intrahepatic Posto Systemic Shunt | EHCF002, EHAF004, EHPF001, EHNF001 |
| Transjugular Liver Biopsy | HLHH001, HLHH005 |
| Transplant Recipient without Liver | Z94.0, Z94.1, Z94.2, Z94.3, Z94.8, Z94.8, T86.1, T86.2, T86.3, T86.8, T86.8, T86.8, Z94801, Z94802, Z94803, Z94804, Z94809, T86.0, JAEA003 |
| Type 1 Diabetes Mellitus | E10- |
| Type 2 Diabetes Mellitus | E11- |
| Urinary Tract Infection | N10-, N39.0 |
| Vasopressor | EQLF001, EQLF003 |

**Table S2: Characteristics of patients hospitalized for hepatitis A according to severity and case-fatality. France, 2013-2024 (n=7,928 patients; univariate analysis)**

| Characteristic | Overall | | Severe Hepatitis A | | | | | Hepatitis A-related death | | | | |
|---|---|---|---|---|---|---|---|---|---|---|---|---|
| | | | No | | Yes | | | No | | Yes | | |
| | n | % | n | % | n | % | | n | % | n | % | |
| | 7,928 | 100 | 5,621 | 70.9 | 2,307 | 29.1 | **p-value[1]** | 7,815 | 98.6 | 113 | 1.43 | **p-value[2]** |
| **Age, Median (IQR)** | 30.0 (18.0, 48.0) | | 28.0 (16.0, 47.0) | | 35.0 (23.0, 51.0) | | <0.001 | 30.0 (18.0, 48.0) | | 70.0 (31.0, 84.0) | | <0.001 |
| **Male Sex** | 4,807 | 60.63 | 3,249 | 57.80 | 1,558 | 67.53 | <0.001 | 4,741 | 60.67 | 66 | 58.41 | 0.63 |
| **Smoking Habits** | 635 | 8.01 | 397 | 7.06 | 238 | 10.32 | <0.001 | 620 | 7.93 | 15 | 13.27 | 0.052 |
| **Alcohol Use Disorders** | 451 | 5.69 | 270 | 4.80 | 181 | 7.85 | <0.001 | 430 | 5.50 | 21 | 18.58 | <0.001 |
| **Obesity** | 626 | 7.90 | 431 | 7.67 | 195 | 8.45 | 0.24 | 605 | 7.74 | 21 | 18.58 | <0.001 |
| **Type-2 Diabetes Mellitus** | 385 | 4.86 | 247 | 4.39 | 138 | 5.98 | 0.003 | 364 | 4.66 | 21 | 18.58 | <0.001 |
| **Liver Risk Factors** | 326 | 4.11 | 201 | 3.58 | 125 | 5.42 | <0.001 | 318 | 4.07 | 8 | 7.08 | 0.14 |
| **Cirrhosis** | 176 | 2.22 | 95 | 1.69 | 81 | 3.51 | <0.001 | 158 | 2.02 | 18 | 15.93 | <0.001 |
| **HIV Infection** | 308 | 3.88 | 189 | 3.36 | 119 | 5.16 | <0.001 | 306 | 3.92 | 2 | 1.77 | 0.33 |
| **CCI ≥ 3** | 667 | 8.41 | 466 | 8.29 | 201 | 8.71 | 0.54 | 595 | 7.61 | 72 | 63.72 | <0.001 |
| **Fdep ≥ Q4** | 3,185 | 41.27 | 2,305 | 42.15 | 880 | 39.13 | 0.014 | 3,130 | 41.15 | 55 | 49.55 | 0.081 |
| **Period** | | | | | | | | | | | | |
| **A** | 2,109 | 27 | 1,619 | 29 | 490 | 21 | | 2,084 | 27 | 25 | 22 | |
| **B** | 2,418 | 30 | 1,598 | 28 | 820 | 36 | | 2,401 | 31 | 17 | 15 | |
| **C** | 1,350 | 17 | 948 | 17 | 402 | 17 | <0.001 | 1,335 | 17 | 15 | 13 | <0.001 |
| **D** | 414 | 5.2 | 303 | 5.4 | 111 | 4.8 | | 400 | 5.1 | 14 | 12 | |
| **E** | 1,637 | 21 | 1,153 | 21 | 484 | 21 | | 1,595 | 20 | 42 | 37 | |

*Severe HA was hepatic and/or extrahepatic organ failure-within 12 weeks of admission with the first record of HA as primary discharge diagnosis. HA-related death was death within 12 weeks post-admission. Periods were A) before the outbreak among MSM; B) MSM outbreak; C) after outbreak until COVID-19 pandemic; D) COVID-19 pandemic, as defined for France; and E) after COVID-19 pandemic.*

*[1] Wilcoxon rank sum test; Pearson's Chi-squared test;*

*[2] Wilcoxon rank sum test; Fisher's Exact Test for Count Data with simulated p-value (based on 2000 replicates)*

**Table S3: Likelihood of Severe Hepatitis A and Hepatitis A-related Death in Propensity-Matched Patients in hospitals, France, 2013-2024**

| Characteristic | Severe HA OR (95% CI)[1] | p-value | HA-related death OR (95% CI)[1] | p-value |
|---|---|---|---|---|
| **Age** | 1.00 (1.00 to 1.00) | >0.99 | 0.99 (0.98 to 1.00) | 0.12 |
| **Male Sex** | 0.98 (0.88 to 1.10) | 0.75 | 0.88 (0.58 to 1.33) | 0.53 |
| **Smoking Habits** | 0.97 (0.81 to 1.15) | 0.71 | 0.66 (0.33 to 1.21) | 0.20 |
| **Alcohol Use Disorders** | 0.99 (0.80 to 1.22) | 0.91 | 1.47 (0.74 to 2.77) | 0.25 |
| **Type-2 Diabetes Mellitus** | 1.05 (0.84 to 1.32) | 0.65 | 0.78 (0.43 to 1.34) | 0.39 |
| **Liver Risk Factors** | 0.91 (0.73 to 1.13) | 0.40 | 0.68 (0.26 to 1.50) | 0.38 |
| **Obesity** | 0.87 (0.73 to 1.05) | 0.14 | 1.20 (0.69 to 2.00) | 0.49 |
| **Cirrhosis** | 1.31 (0.96 to 1.77) | 0.087 | 1.02 (0.49 to 2.03) | 0.95 |
| **CCI ≥ 3** | 0.88 (0.71 to 1.10) | 0.26 | 1.25 (0.67 to 2.29) | 0.48 |
| **Fdep Q4 or Q5** | 1.00 (0.91 to 1.11) | 0.93 | 1.09 (0.73 to 1.61) | 0.67 |
| **Period**<br>**A [2013-2017[** | Reference | | Reference | |
| **B [2017-Jun. 2018[** | 1.49 (1.30 to 1.71) | <0.001 | 1.06 (0.55 to 2.02) | 0.86 |
| **C [Jul. 2018-2019]** | 1.54 (1.31 to 1.80) | <0.001 | 1.07 (0.52 to 2.12) | 0.84 |
| **D [2020-Jun. 2021]** | 1.36 (1.06 to 1.74) | 0.014 | 1.75 (0.82 to 3.58) | 0.13 |
| **E [Jul. 2021-2024]** | 1.60 (1.38 to 1.86) | <0.001 | 2.22 (1.32 to 3.84) | 0.003 |

*Risks were assessed using multivariate logistic regression models in propensity-matched samples (see figures S5 and S6 for covariates balances) adjusted for all studied variables.*

*Severe HA was hepatic and/or extrahepatic organ failure-within 12 weeks of admission with the first record of HA as primary discharge diagnosis. HA-related death was death within 12 weeks post-admission.*

*[1] OR = Odds Ratio, CI = Confidence Interval*

**Supplementary Figures**

**Figure S1: Study Flowchart**

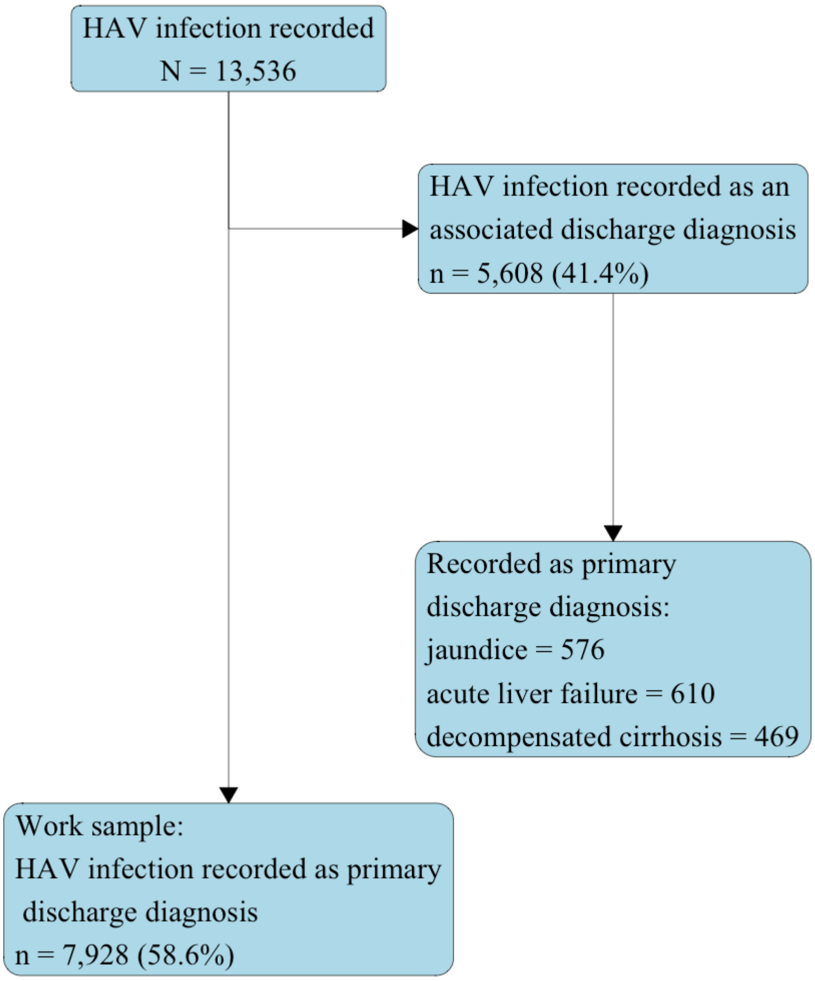


*All patients with a primary or associated discharge diagnosis of hepatitis A (ICD-10 code B150 or B159) between January 2013 and December 2024 were considered. To increase specificity, only patients with Hepatitis A recorded as the primary discharge diagnosis (58.6%) were retained for analysis.*

**Figure S2: Quarterly number of HA cases recorded in hospitals, France, 2013-2024**

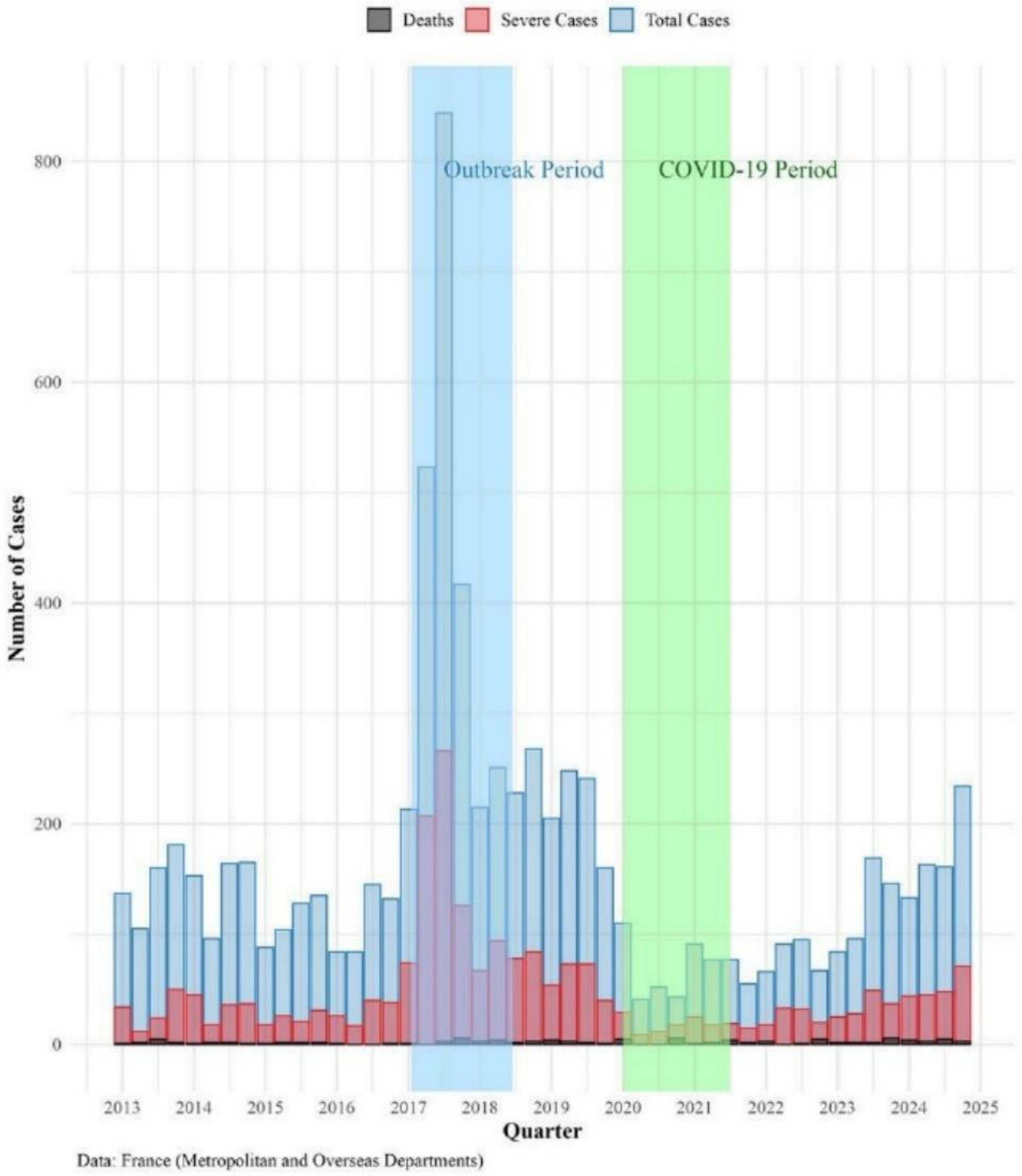


*Total number of HA cases (blue), of severe cases (red) and of HA-related deaths (black) are shown. Two periods are highlighted: the outbreak among MSM (blue) and the COVID-19 pandemic (green).*

**Figure S3: Covariate balance after propensity score matching for severe Hepatitis A (A) and Hepatitis A-Related Death (B), Frech Hospitals, 2013-2024**

*A)*

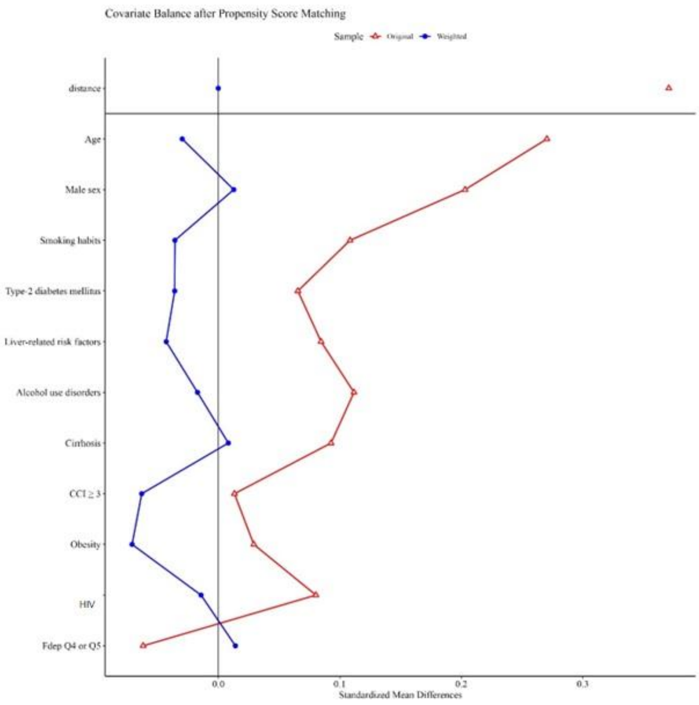


B)

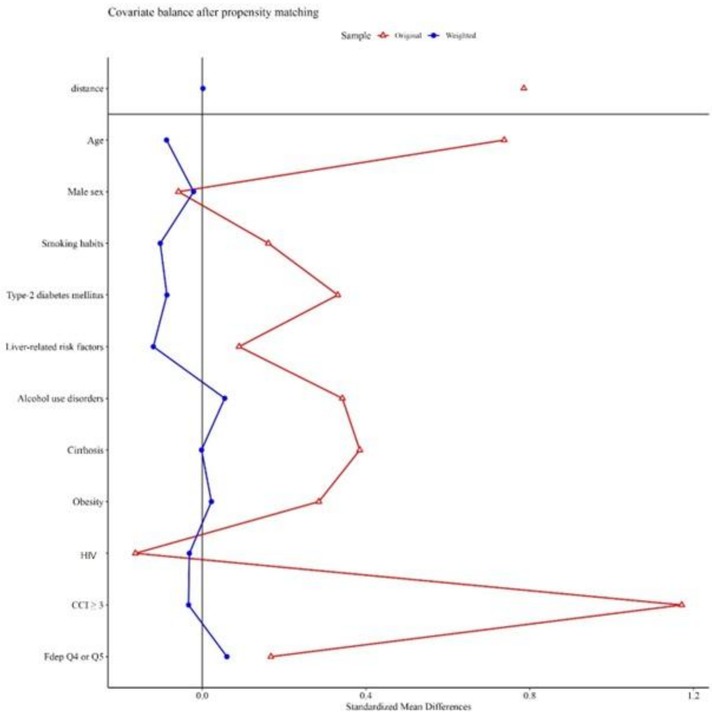


*A full matching algorithm (N = 7651 for A and N = 7717 for B) was applied using the estimated propensity scores.*
